# Hidden in Success: Gendered Patterns of Suboptimal Care Engagement Among TB Patients Who “Successfully” Completed Treatment in South Africa

**DOI:** 10.1101/2025.09.04.25335036

**Authors:** Andrew Medina Marino, Eke Arua, Lindsey de Vos, Kuhle Fiphaza, Dana Bezuidenhout, Nondumiso Ngcelwane, Salome Charalambous, Joseph Daniels

## Abstract

**Background:** Adherence to tuberculosis (TB) treatment is key to optimal health outcomes. Programmatic definitions of treatment success may mask heterogeneity in care engagement patterns that increase risk of unfavorable outcomes.

**Methods:** Using patient-level medication refill data, latent-class growth modelling was used to identify longitudinal trajectories of care engagement among participants who programmatically achieved treatment success. Logistic regression was conducted to investigate participant-level characteristics associated with trajectory class membership.

**Results:** Among 548 participants, we identified three trajectories: Class 1 (consistent engagement; 84.1%), Class 2 (suboptimal engagement after 2 months; 7.7%) and Class 3 (suboptimal engagement from initiation; 8.2%). At treatment completion, Classes 1-3 accumulated 9.7 (95% CI: 7.4–11.8), 68.4 (60.4–76.9) and 55.5 (48.1–62.7) missed refill days, respectively. In gender-stratified models, men exhibited all three trajectories (83.1%, 7.4%, and 9.5%, respectively), and accumulated 10.6 [7.8–13.3], 61.0 [50.2–71.3], 53.3 [53.3–71.4] missed refill days, respectively. Women exhibited only Classes 1 and 3 (89.5% and 10.5%, respectively) and accumulated 12.1 [7.8–16.5] and 46.9 [33.3–61.6] missed refill days, respectively. Among men, prior TB (Class 2: aOR 7.44, 2.79–19.8; Class 3: aOR 2.78, 1.07–7.25) and HIV-negative status (Class 3: aOR 2.72, 1.13–6.54) were associated with suboptimal trajectories. Among women, prior TB was associated with suboptimal engagement (aOR 5.22, 1.11–24.44).

**Conclusion:** Programmatic Treatment Success obscured suboptimal engagement trajectories. Patient-centered counseling and gender-responsive interventions are needed to address suboptimal engagement across treatment stages. Shorter treatment regimens will unlikely resolve suboptimal engagement, underscoring the importance of regimen forgiveness.

**Article Summary:** Among South African TB patients programmatically classified as treatment successes, trajectory modeling revealed hidden suboptimal engagement patterns, particularly among men. Prior TB and HIV-negative status were associated with suboptimal engagement, underscoring the need for gender-responsive, patient-centered counseling and support interventions

## INTRODUCTION

Since 2010, the World Health Organization (WHO) has recommended a 6-month, multi-drug regimen of daily treatment for people diagnosed with drug-susceptible TB (DS-TB).[1] In 2022, among all people who started therapy for DS-TB globally, the treatment success rate was 86%, short of the End TB Strategy target of ≥90% by 2025.[2] In South Africa, one of the highest TB burden countries, the treatment success rate was 83% in 2023.[3] Treatment success is a cornerstone metric of TB programs globally. However, its current definition is centered on whether a patient ultimately collects and finishes the prescribed regimen without a treatment interruption of ≥60 consecutive days (i.e., lost to follow-up).[4] This fails to account for the timing (e.g., intensive vs continuation phase), pattern (i.e., clustered vs dispersed) and cumulative duration of missed treatment. Consequently, patients classified as completing treatment may experience fragmented or protracted courses of care with varying engagement and adherence patterns, but remain invisible in aggregate program indicators for being at increased risk of morbidity, mortality and post-TB sequelae.[5–7]

Evidence from both trial and programmatic settings underscores the limited tolerance for missed TB doses. In a pooled patient-level meta-analysis of treatment-shortening trials, adherence was the strongest predictor of success. Specifically, in both the 4-month experimental and 6-month control regimens, patients who missed ≥10% of doses had roughly a six-fold higher hazard of unfavorable outcomes (i.e., failure, relapse, death) compared to those who were fully adherent (7 of 7 doses/week).[8] A multicenter cohort study from Brazil conducted under routine programmatic conditions found a similar gradient, with ≥10% missed doses associated with an 81% probability of unfavorable outcome, and each 1% of non-adherence increasing the odds by 11%.[9] Evidence also suggests that the timing and distribution of missed doses may drive risk. Specifically, a pooled analysis of individual-level data from three clinical trials reported that missing four treatment days in a month increased the hazard of unfavorable outcomes by 61%, with clustered gaps posing greater risk than the same number of missed doses dispersed over time.[7] These findings highlight the limited margin for missed doses in standard 6-month regimens, suggesting that even among patients who complete treatment by programmatic definitions, intermittent missed dosing due to gap in picking up treatment refills may carry substantial clinical risk.

Despite growing investment in behavioral interventions to improve TB treatment outcomes, many are designed with a one-size-fits-all approach, assuming uniform patient behavior and support needs.[10,11] This overlooks important variation in how individuals engage with care and the structural, social, gendered and individual factors influencing that engagement.[12,13] As evidence mounts that the pattern of missed doses, not just the amount, shapes treatment outcomes,[7] identifying patterns and factors associated with care engagement is crucial to implementing targeted, differentiated strategies that support sustained engagement in care; this may be of unique importance among those that would otherwise be identified programmatically as successfully completing treatment.[14] Towards this, we conducted latent class trajectory modelling using treatment refill data to identify patterns of care engagement among men and women with DS-TB programmatically classified as successfully completing treatment, and identified factors associated with class membership.

## METHODS

### Study Design and Setting

A prospective cohort study was conducted from February 2021 through August 2022 in 21 government healthcare facilities across Buffalo City Metro (BCM) Health District, Eastern Cape Province, South Africa. In 2023, Eastern Cape had the highest provincial TB notification rate in South Africa (703 per 100,000 population), with BCM having a TB notification rate of between 900-999 cases per 100,000 population.[15] That same year, Eastern Cape had an estimated general HIV prevalence of 12.7% (95% CI: 12.2%-13.3%).[16] In 2022, BCM had a DS-TB treatment success rate of 67.4%; the national treatment success rate was 75.8%.[17]

### Participant Recruitment

Individuals initiating or already engaged in TB treatment were screened for eligibility: 1) aged ≥18 years, 2) fluent in English or isiXhosa, 3) residency within a catchment area of one of the 21 collaborating health facilities, and 4) provision of informed consent. Individuals with extrapulmonary TB without lung involvement or drug-resistant TB were excluded. Eligible individuals providing written consent were provided R50 (∼$3.33USD) and a small snack for their time.

### Data Collection and Study Measures

After consent, participants completed a research-staff administered questionnaire which included questions on socio-demographic factors, health characteristics,[18] health behaviors,[19] TB knowledge,[20] attitudes and beliefs [21–23] using validated, locally adapted measures. Questionnaires were developed in English, translated into isiXhosa, and back translated to English to ensure accuracy. Care engagement history—including treatment start date, dates of expected and actual medication refill visits, and treatment outcomes—was extracted from participants’ medical records.

### Outcome Definition

The primary outcome was the longitudinal trajectories of cumulative missed TB medication refill days since treatment initiation among those classified as “Treatment Success” (i.e., composite of Treatment Completed and Cured) per WHO guidelines. Missed refill days were calculated as the difference in days between a participant’s scheduled medication refill date and the actual date they presented to refill their medication. Participants who refilled their medication prior to the scheduled refill date were not considered to have missed a day. While medication refill timeliness can serve as a proxy for adherence when direct measures of adherence are unavailable,[24] here it is used primarily as an indicator of engagement in care, reflecting patients’ continued and timely interaction with health services rather than direct evidence of daily medication-taking.

### Statistical Analysis

Our analytical sample included participants programmatically classified as Treatment Success. Participant characteristics were summarized as medians (IQR) for continuous variables and frequencies (%) for categorical variables, with comparisons by Wilcoxon rank-sum, chi-square, or Fisher’s exact tests as appropriate.

Latent-class growth modelling was used to identify trajectories of total missed TB medication refill days from treatment initiation through the end of the expected 6-month treatment period. Linear, quadratic, and cubic models were explored, with the latter two offering greater flexibility in capturing the shape of the trajectories. The latent class growth models with a minimum of two classes were fit to the analytic dataset, and separately by gender. Model fit was evaluated using Bayesian information criteria (BIC), Akaike information criteria (AIC), and entropy, with class size and subject-matter expertise guiding final model selection. Participants were assigned to classes based on the highest predicted probability of membership.

Multinomial logistic regression was used to investigate participants’ characteristics associated with trajectory group membership in the overall and gender-stratified datasets; age, gender and education level were included as potential confounders. Missing covariate data were handled using multiple imputation. Analyses were conducted in **R** (version 4.4.2), using the **lcmm** package for latent-class growth modelling.[25,26] Adjusted Odds Ratios (aOR) are reported.

### Ethical Considerations

Ethics approval was obtained from the Human Research Ethics Committee of the Faculty of Health Sciences, University of Cape Town (Ref no.: 673/2019) with an institutional reliance agreement by Arizona State University. Study approval was provided by the Eastern Cape Provincial Department of Health (Ref no.: EC202010_023). All participants provided written informed consent.

## RESULTS

Of 657 enrolled participants, 548 (83.4%) were programmatically classified as Treatment Success and included in the analytic dataset (Table 1); of note, 72/657 (11%) did not complete treatment (i.e., composite of Loss-to-Follow Up, Died and Treatment Failure), and 37/657 (5.6%) did not have a treatment outcome assigned. Among those, the median age was 38 years (IQR: 30–47), most were men (67%), unemployed (78.3%), had less than a high school qualification (94.5%), and reported low levels of alcohol use (86.9%). Overall, 46.2% self-reported living with HIV, 28.1% had a previous history of TB, 38.9% screened positive for moderate to severe depression, and 27.7% reported high levels of medical mistrust. Compared to women, men were more likely to live alone (23.2% vs. 14.4%; p=0.02), have less education (p=0.02) and moderately higher levels of TB knowledge (p=0.07). In comparison, women were more likely to live with children (74.0% vs. 55.0%; p<0.01), be living with HIV (63.0% vs. 37.9%; p<0.01), and report higher anxiety scores (p=0.024).

**Table 1:** Characteristics of study participants stratified by gender. (All data are presented as N (%) unless stated otherwise)

| Characteristics | Overall<br>(N=548) | Men<br>(n=367; 67%) | Women<br>(n=181; 33%) | p-value |
| --- | --- | --- | --- | --- |
| **Socio-Demographic Characteristics** |  |  |  |  |
| **Age [Median (IQR)]** | 38 (30, 47) | 39 (31, 47) | 38 (30, 48) | 0.6 |
| **Relationship Status** |  |  |  | 0.8 |
| Not in a relationship | 375 (68.4) | 250 (68.1) | 125 (69.1) |  |
| In a relationship | 173 (31.6) | 117 (31.9) | 56 (30.9) |  |
| **Level of Education** |  |  |  | 0.02 |
| Primary and below | 109 (19.9) | 82 (22.3) | 27 (14.9) |  |
| Secondary (before Matric<sup>a</sup>) | 409 (74.6) | 270 (73.6) | 139 (76.8) |  |
| Matric<sup>a</sup> and above | 30 (5.5) | 15 (4.1) | 15 (8.3) |  |
| **Lives Alone** |  |  |  | 0.02 |
| Yes | 111 (20.3) | 85 (23.2) | 26 (14.4) |  |
| No | 437 (79.7) | 282 (76.8) | 155 (85.6) |  |
| **Lives with Children** |  |  |  | <0.01 |
| Yes | 336 (61.3) | 202 (55.0) | 134 (74.0) |  |
| No | 103 (18.8) | 82 (22.3) | 21 (11.6) |  |
| Missing | 109 (19.9) | 83 (22.6) | 26 (14.4) |  |
| **Employment Status** |  |  |  | 0.30 |
| Employed | 119 (21.7) | 84 (22.9) | 35 (19.3) |  |
| Unemployed | 429 (78.3) | 283 (77.1) | 146 (80.7) |  |
| **Monthly Household Income (Rands)** |  |  |  | 0.40 |
| <R2000 | 368 (67.2) | 251 (68.4) | 117 (64.6) |  |
| R2000 - R5000 | 139 (25.4) | 87 (23.7) | 52 (28.7) |  |
| >R5000 | 41 (7.5) | 29 (7.9) | 12 (6.6) |  |
| **Social Support** |  |  |  | 0.9 |
| Low (<3) | 24 (4.4) | 16 (4.4) | 8 (4.4) |  |
| Moderate (4-5) | 175 (31.9) | 122 (33.2) | 53 (29.3) |  |
| High (>5) | 342 (62.4) | 224 (61.0) | 118 (65.2) |  |
| Missing | 7 (1.3) | 5 (1.4) | 2 (1.1) |  |
| **Social Capital** |  |  |  | 0.4 |
| Low (0-3) | 61 (11.1) | 38 (10.4) | 23 (12.7) |  |
| Med (4-6) | 370 (67.5) | 244 (66.5) | 126 (69.6) |  |
| High (7-9) | 112 (20.4) | 80 (21.8) | 32 (17.7) |  |
| Missing | 5 (0.9) | 5 (1.4) | 0 (0) |  |
| **Clinical Characteristics and Health Behaviors** |  |  |  |  |
| **Ever had TB Before** |  |  |  | 0.2 |
| Never | 394 (71.9) | 258 (70.3) | 136 (75.1) |  |
| Yes, less than 2 years ago | 50 (9.1) | 40 (10.9) | 10 (5.5) |  |
| Yes, more than 2 years ago | 104 (19.0) | 69 (18.8) | 35 (19.3) |  |
| **HIV Status** |  |  |  | <0.01 |
| Positive | 253 (46.2) | 139 (37.9) | 114 (63.0) |  |
| Negative | 270 (49.3) | 210 (57.2) | 60 (33.1) |  |
| Unknown | 25 (4.6) | 18 (4.9) | 7 (3.9) |  |
| **Depression (PHQ-9)<sup>b</sup>** |  |  |  | 0.3 |
| None/Minimal (0-4) | 243 (44.3) | 170 (46.3) | 73 (40.3) |  |
| Mild (5-6) | 87 (15.9) | 57 (15.5) | 30 (16.6) |  |
| Moderate (7-14) | 160 (29.2) | 102 (27.8) | 58 (32.0) |  |
| Moderate-Severe (≥15) | 53 (9.7) | 33 (9.0) | 20 (11.0) |  |
| Missing | 5 (0.9) | 5 (1.4) | 0 (0) |  |
| **Anxiety (GAD-7)** |  |  |  | 0.024 |
| Minimal (0-4) | 338 (61.7) | 234 (66.8) | 104 (57.5) |  |
| Mild (5-9) | 142 (25.9) | 94 (25.7) | 48 (26.5) |  |
| Moderate (10-14) | 43 (7.8) | 24 (6.5) | 19 (10.5) |  |
| Severe (15-21) | 20 (3.6) | 10 (2.7) | 10 (5.5) |  |
| Missing | 5 (0.9) | 5 (1.4) | 0 (0) |  |
| **Alcohol Use (AUDIT)** |  |  |  | 0.4 |
| Low (0-7) | 476 (86.9) | 312 (85.0) | 164 (90.6) |  |
| Medium (8-15) | 45 (8.2) | 34 (9.3) | 11 (6.1) |  |
| High (16-20) | 11 (2.0) | 8 (2.2) | 3 (1.7) |  |
| Alcohol Dependent (>20) | 9 (1.6) | 8 (2.2) | 1 (0.6) |  |
| Missing | 7 (1.3) | 5 (1.4) | 2 (1.1) |  |
| **Knowledge, Attitudes and Beliefs** |  |  |  |  |
| **TB Knowledge** |  |  |  | **0.07** |
| Low | 257 (46.9) | 162 (44.1) | 95 (52.5) |  |
| High | 286 (52.2) | 200 (54.5) | 86 (47.5) |  |
| Missing | 5 (0.9) | 5 (1.4) | 0 (0) |  |
| **HIV Stigma [Median (IQR)]** | 16 (11, 20) | 16 (11, 20) | 16 (12, 21) | 0.4 |
| **TB Stigma [Median (IQR)]** |  |  |  |  |
| Disclosure | 8 (5, 10) | 8 (5, 10) | 9 (5, 10) | 0.9 |
| Isolation | 6 (4, 8) | 6 (4, 8) | 6 (4, 8) | 0.8 |
| **Medical Mistrust** |  |  |  | 0.4 |
| Low (1-19) | 152 (27.7) | 101 (27.5) | 51 (28.2) |  |
| Medium (20-37) | 209 (38.1) | 134 (36.5) | 75 (41.4) |  |
| High (38-55) | 141 (25.7) | 100 (27.2) | 41 (22.7) |  |
| Missing | 46 (8.4) | 32 (8.7) | 14 (7.7) |  |

<sup>a</sup> Matric is the equivalent of a high school certificate which is awarded at the end of secondary school.
<sup>b</sup> Score Ranges Based on a locally validated PHQ-9.

### Overall Cohort Trajectory Model

The quadratic model with three distinct trajectory classes provided the best fit for the overall cohort (Figure 1; Supplementary Table 1). Classes were descriptively named based on participants’ patterns of engagement: Class 1 (consistent) was characterized by slow, small increases in the cumulative number of days missed for treatment refill pickups over the six-month treatment period; Class 2 (suboptimal after 2 months) was characterized by a steep increase in the cumulative number of days missed for treatment refill pickups approximately two months following treatment initiation; Class 3 (suboptimal from onset) was characterized by an immediate, steep increase in the cumulative number of days missed for treatment refill pickups from the start of treatment. Overall, 84.1% of the overall cohort were members of Class 1, 7.7% were members of Class 2 and 8.2% were members of Class 3. Participants allocated to Classes 1, 2 and 3 were predicted to miss their treatment refill dates by an average of 0.84 (95% CI: 0–1.89), 3.49 (95% CI: 0–6.85) and 7.49 (95% CI: 3.94–10.87) days, respectively. At the time of treatment completion, participants allocated to Classes 1, 2 and 3 were predicted to accumulate, on average, a total of 9.68 (95% CI: 7.41–11.83), 68.42 (95% CI: 60.35–76.92) and 55.47 (95% CI: 48.05–62.66) days of missed treatment refill dates, respectively.

**Figure 1.**
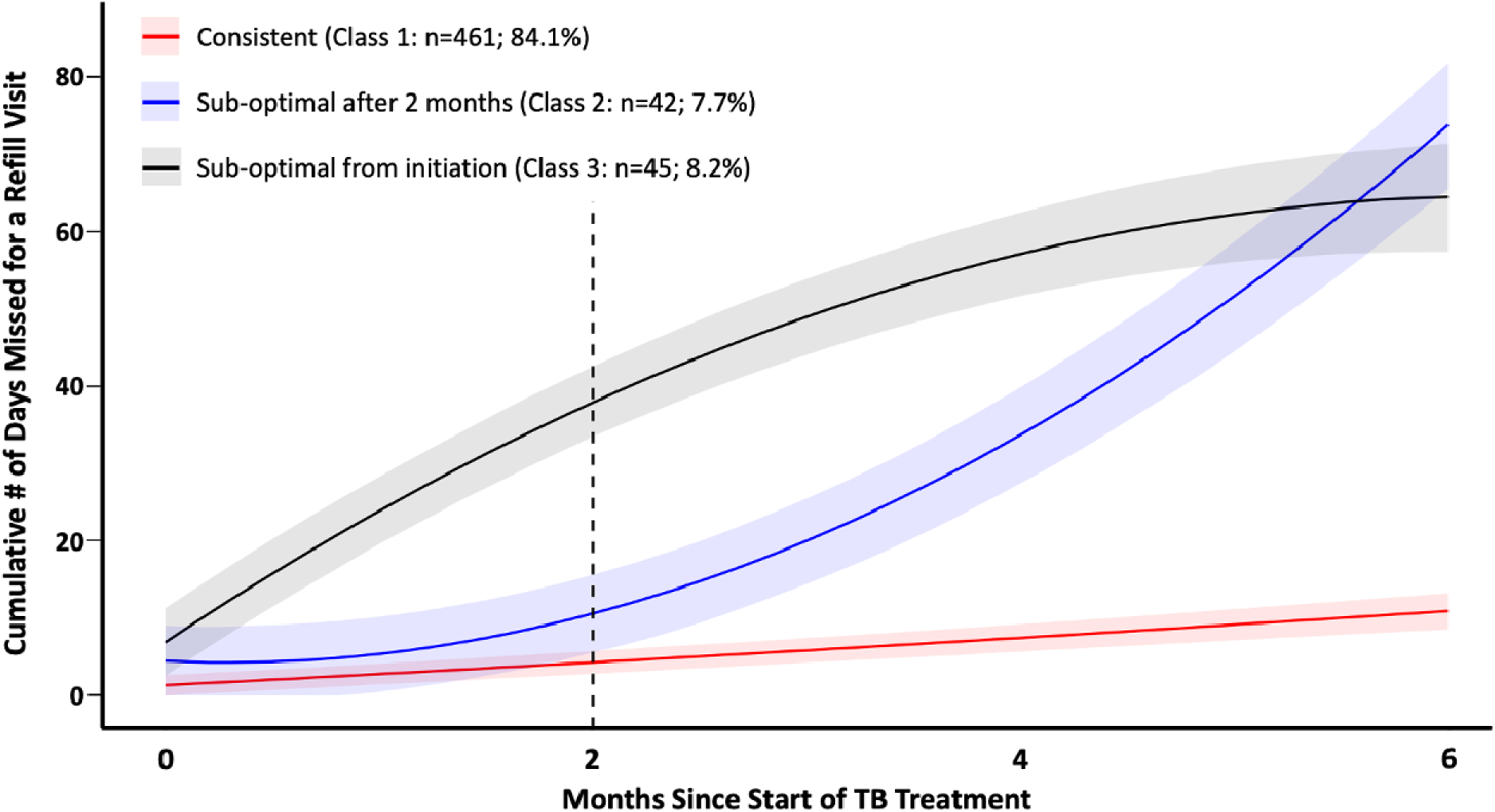
Latent class trajectories of cumulative number of missed refill days among all study participants (N=548). Predicted mean trajectories (curved lines) and 95% confidence intervals (shaded area). Dotted vertical line at 2-month period represents the end of intensive phase of TB treatment.

### Gender Specific Trajectory Models

Modelling revealed three trajectory classes for men and two for women (Figure 2; Supplementary Table 1). Men’s trajectory classes mirrored those for the overall cohort in size, trajectory, and description (i.e., Class 1: consistent (83.1%); Class 2: suboptimal after 2 months (7.4%%); Class 3: suboptimal from onset (9.5%)). In comparison, women’s trajectory classes mirrored the overall cohort’s consistent (Class 1; 89.5%) and suboptimal from onset (Class 3; 10.5%) groups. Though most men and women were assigned to their respective consistent class trajectories, a smaller proportion of men were assigned to this class compared to women (83.1% vs 89.5%, respectively). In comparison, the proportions of men and women assigned to their respective suboptimal from onset trajectories (Class 3) were similar (9.5% vs 10.5%, respectively). The difference between the proportion of men and women with overall suboptimal engagement (i.e., combined Class 2 and 3: Men=16.9% vs Women=10.5%) was driven by the 7.4% of men in the suboptimal after 2 months group. Men assigned to Classes 1, 2 and 3 were predicted to miss their treatment refill dates by an average of 0.83 (95% CI: 0–2.37), 4.19 (95% CI: 0–9.34) and 8.31 (95% CI:3.69–13.13) days, respectively, and accumulate a total average of 10.57 (95% CI: 7.78–13.29), 61.03 (50.15–71.32) and 53.30 (53.30–71.35) days’ worth of treatment refill dates by the time of treatment completion. In comparison, women assigned to Classes 1 and 3 were predicted to miss their treatment refill dates by an average of 1.02 (95% CI: 0.27–1.79) and 1.81 (95% CI: 0–4.22) days, respectively, and accumulated a total average of 12.12 (95% CI: 7.84–16.46) and 46.94 (95% CI: 33.27–61.62) days’ worth of treatment refill dates at the time of treatment completion.

**Figure 2.**
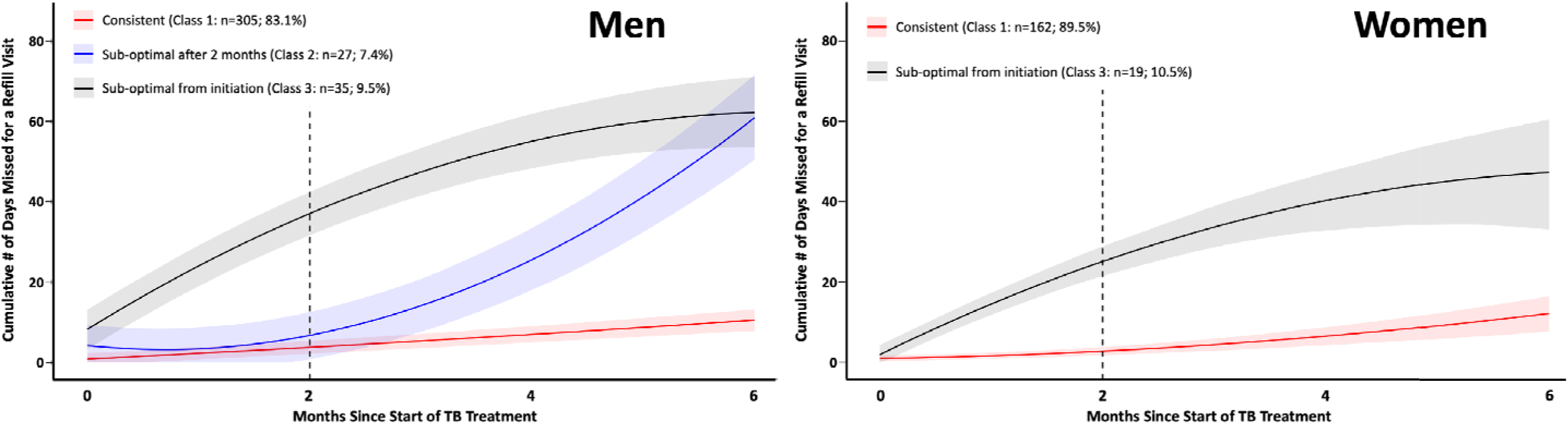
Latent class trajectories of cumulative number of missed refill days for men (n=367; left) and women (n=181; right). Predicted me (curved lines) and 95% confidence intervals (shaded area). Dotted vertical line at 2-month period represents the end of intensive phase of TB treatment

### Characteristics Associated with Trajectory Class Membership: Overall Cohort

The consistent engagement group (Class 1) was used as the reference group, with associations reported for the suboptimal after 2 months (Class 2) and suboptimal from initiation (Class 3). Additionally, classes 2 and 3 were combined to explore which variables may be associated with “Overall Suboptimal Engagement” in care. For the overall cohort, having previously had TB within the past two years had a positive and significant association with suboptimal after 2 months (aOR 5.94, 95% CI: 2.58–13.68), suboptimal from initiation (aOR 3.34, 95% CI: 1.43–7.81) and overall suboptimal engagement (aOR 4.38, 95% CI: 2.29–8.36) (Table 2). Descriptive statistics are reported in Table S2.

**Table 2.** Logistic regression analysis of associations between participant characteristics and assigned trajectory classes, adjusted for age, gender and education level, among all participants.

|  | Suboptimal Engagement after 2 months<sup>a</sup> |  | Suboptimal Engagement from Initiation<sup>a</sup> |  | Overall Suboptimal Engagement<sup>b</sup> |  |
| --- | --- | --- | --- | --- | --- | --- |
|  | aOR | p-value | aOR | p-value | p-value | p-value |
| **Relationship Status** |  |  |  |  |  |  |
| In a relationship | 0.65 (0.31-1.36) | 0.25 | 0.62 (0.29-1.34) | 0.22 | 0.63 (0.37–1.10) | 0.10 |
| **Lives Alone** |  |  |  |  |  |  |
| No | 2.74 (0.94-7.97) | 0.06 | 1.37 (0.61-3.07) | 0.45 | 0.93 (0.58–1.48) | 0.75 |
| **Live with Children** |  |  |  |  |  |  |
| Yes | 1.77 (0.81-3.88) | 0.15 | 1.65 (0.74-3.68) | 0.22 | 1.06 (0.61-1.84) | 0.69 |
| **Employment Status** |  |  |  |  |  |  |
| Unemployed | 0.72 (0.34-1.51) | 0.39 | 1.04 (0.48-2.25) | 0.92 | 0.87 (0.50–1.52) | 0.62 |
| **Monthly Household Income (ref=<R2000)** |  |  |  |  |  |  |
| R2000 - R5000 | 1.38 (0.68-2.79) | 0.38 | 1.31 (0.65-2.64) | 0.44 | 1.37 (0.81-2.30) | 0.24 |
| >R5000 | 0.74 (0.16-3.50) | 0.70 | 1.20 (0.34-4.27) | 0.78 | 0.98 (0.35–2.72) | 0.97 |
| **Social Support (ref=High)** |  |  |  |  |  |  |
| Low | 0.62 (0.08-4.90) | 0.65 | 0.50 (0.06-3.90) | 0.51 | 0.55 (0.12–2.46) | 0.44 |
| Moderate | 1.33 (0.69-2.58) | 0.39 | 0.98 (0.51-1.90) | 0.96 | 1.14 (0.70–1.85) | 0.61 |
| **Ever had TB (ref=Never)** |  |  |  |  |  |  |
| Yes, less than 2 years ago | **5.94 (2.58-13.68)** | **<0.01** | **3.34 (1.43-7.81)** | **<0.01** | **4.38 (2.29–8.36)** | **<0.01** |
| Yes, more than 2 years ago | 1.27 (0.55-2.97) | 0.57 | 1.12 (0.49-2.57) | 0.79 | 1.18 (0.64–2.18) | 0.60 |
| **HIV Status (ref=Positive)** |  |  |  |  |  |  |
| Negative | 1.22 (0.62-2.40) | 0.56 | 1.56 (0.79-3.08) | 0.20 | 1.41 (0.86–2.32) | 0.17 |
| Unknown | 1.11 (0.24-5.18) | 0.89 | 1.25 (0.27-5.90) | 0.76 | 1.19 (0.38–3.70) | 0.77 |
| **Depression (ref=Minimal; 0-4)** |  |  |  |  |  |  |
| Mild | 1.28 (0.56-2.95) | 0.56 | 1.12 (0.45-2.79) | 0.81 | 1.02 (0.93-1.12) | 0.57 |
| Moderate | 0.73 (0.33-1.61) | 0.43 | 1.40 (0.69-2.83) | 0.62 | 1.04 (0.61–1.78) | 0.88 |
| Moderate-Severe | 0.43 (0.10-1.91) | 0.27 | 1.02 (0.33-3.16) | 0.97 | 0.71 (0.29–1.78) | 0.47 |
| **Anxiety (ref=Minimal: 0-4)** |  |  |  |  |  |  |
| Mild | 1.56 (0.78-3.12) | 0.20 | 1.08 (0.52-2.21) | 0.84 | 1.31 (0.78–2.20) | 0.31 |
| Moderate | 0.64 (0.14-2.85) | 0.56 | 1.12 (0.37-3.42) | 0.84 | 0.89 (0.35–2.23) | 0.80 |
| Severe | 0.69 (0.09-5.52) | 0.73 | 0.58 (0.07-4.58) | 0.61 | 0.63 (0.14–2.83) | 0.55 |
| **Alcohol Use (ref = Low)** |  |  |  |  |  |  |
| Medium | 1.75 (0.63–4.87) | 0.28 | 1.69 (0.71–4.05) | 0.23 | 1.82 (0.87–3.81) | 0.12 |
| High/Addiction | 0.58 (0.07–4.57) | 0.61 | 0.58 (0.07–4.50) | 0.60 | 0.58 (0.13–2.55) | 0.46 |
| **TB Knowledge** |  |  |  |  |  |  |
| High | 0.93 (0.49–1.78) | 0.83 | 0.93 (0.50–1.74) | 0.82 | 0.93 (0.58–1.48) | 0.75 |
| **HIV Stigma [Median (IQR)]** | 0.98 (0.94-1.02) | 0.27 | 1.04 (0.97-1.09) | 0.06 | 1.01 (0.98–1.04) | 0.57 |
| **TB Stigma [Median (IQR)]** |  |  |  |  |  |  |
| Isolation | 0.96 (0.86-1.08) | 0.49 | 1.03 (0.92-1.16) | 0.56 | 1.00 (0.92–1.08) | 0.57 |
| Disclosure | 0.96 (0.88-1.05) | 0.89 | 1.06 (0.97-1.17) | 0.20 | 1.01 (0.94-1.08) | 0.80 |
| **Medical Mistrust (ref=Low)** |  |  |  |  |  |  |
| Medium | 0.93 (0.41-2.13) | 0.87 | 1.43 (0.67-3.06) | 0.35 | 1.18 (0.65–2.14) | 0.58 |
| High | 1.16 (0.49-2.72) | 0.74 | 1.10 (0.46-2.64) | 0.82 | 1.07 (0.57–2.02) | 0.84 |

<sup>a</sup> Odds ratios are from a multinomial logistic regression
<sup>b</sup> Odds ratios are from a logistic regression

### Characteristics Associated with Trajectory Class Membership: Gender Specific Cohorts

For both men and women, their respective consistent engagement groups (Class 1) were used as the reference, with associations reported for the suboptimal after 2 months (Class 2), suboptimal from initiation (Class 3), and a combined “Overall Suboptimal Engagement” for men, and the suboptimal from initiation (Class 3) for women.

Among men, characteristics with a positive and significant association with suboptimal engagement trajectories included: 1) having previously had TB within the past two years (suboptimal after 2 months: aOR 7.44, 95% CI: 2.79–19.8; suboptimal from initiation: aOR 2.78, 95% CI: 1.07–7.25; overall suboptimal: aOR 3.77, 95% CI: 1.82–7.78), and 2) being HIV negative (suboptimal from initiation: aOR 2.72, 95% CI: 1.13–6.54; overall suboptimal: aOR 1.16, 95% CI: 1.02–1.32) (Table 3). Among women, having had TB in the last two years had a positive and significant association with suboptimal from initiation (aOR 5.22, 95% CI: 1.11–24.44), while having had TB more than two years ago had a positive, but statically moderate association with suboptimal from initiation (aOR 2.99, 95% CI: 0.06–9.32) (Table 4). Descriptive statistics are reported in Tables S3 and S4 for men and women, respectively.

**Table 3:** Logistic regression analysis of associations between male participants’ characteristics and assigned trajectory groups or overall suboptimal engagement, adjusted for age and education level.

| Characteristics | Suboptimal Engagement after 2 months<sup>a</sup> |  | Suboptimal Engagement from Initiation<sup>a</sup> |  | Suboptimal Engagement (Collapsed Groups)<sup>b</sup> |  |
| --- | --- | --- | --- | --- | --- | --- |
|  | aOR | p-value | aOR | p-value | aOR | p-value |
| **Relationship Status** |  |  |  |  |  |  |
| In a relationship | 0.46 (0.17–1.25) | 0.12 | 0.72 (0.30–1.69) | 0.45 | 0.91 (0.77-1.06) | 0.23 |
| **Lives Alone** |  |  |  |  |  |  |
| No | 1.88 (0.62–5.69) | 0.26 | 1.57 (0.62–3.98) | 0.34 | 1.10 (0.94-1.29) | 0.48 |
| **Live with Children** |  |  |  |  |  |  |
| Yes | 1.70 (0.70–4.15) | 0.26 | 2.49 (0.79–7.84) | 0.11 | 1.42 (0.73-2.77) | 0.30 |
| **Employment Status** |  |  |  |  |  |  |
| Unemployed | 0.86 (0.34–2.16) | 0.75 | 0.94 (0.41–2.20) | 0.90 | 0.98 (0.84-1.15) | 0.82 |
| **Monthly Household Income (ref=<R2000)** |  |  |  |  |  |  |
| R2000-R5000 | 0.97 (0.37–2.54) | 0.94 | 1.52 (0.70–3.33) | 0.29 | 1.07 (0.92-1.26) | 0.30 |
| >R5000 | 0.95 (0.19–4.78) | 0.95 | 0.95 (0.20–4.44) | 0.95 | 0.99 (0.77-1.28) | 0.96 |
| **Social Support** |  |  |  |  |  |  |
| Low | 1.10 (0.13–9.21) | 0.93 | 0.00 (0.00–Inf) | 0.99 | 0.5 (0.1-2.68) | 0.42 |
| Moderate | 1.57 (0.69–3.58) | 0.29 | 0.94 (0.44–1.99) | 0.87 | 1.35 (0.78-2.33) | 0.28 |
| **Ever had TB (ref=Never)** |  |  |  |  |  |  |
| Yes, less than 2 years ago | **7.44 (2.79–19.8)** | **<0.01** | **2.78 (1.07–7.25)** | **<0.01** | **3.77 (1.82-7.78)** | **<0.01** |
| Yes, more than 2 years ago | 1.79 (0.64–5.05) | 0.27 | 0.61 (0.20–1.84) | 0.38 | 1.41 (0.74-2.69) | 0.30 |
| **HIV Status (ref=Positive)** |  |  |  |  |  |  |
| Negative | 1.15 (0.50–2.68) | 0.74 | **2.72 (1.13–6.54)** | **0.03** | **1.16 (1.02-1.32)** | **0.03** |
| Unknown | 1.78 (0.35–9.03) | 0.49 | 2.56 (0.48–13.72) | 0.27 | 1.17 (0.86-1.60) | 0.30 |
| **Depression (ref=Minimal: 0-4)** |  |  |  |  |  |  |
| Mild | 1.48 (0.30–7.25) | 0.62 | 1.50 (0.61-3.71) | 0.38 | 1.21 (0.51-2.91) | 0.66 |
| Moderate | 0.70 (0.27–1.86) | 0.48 | 1.23 (0.55–2.76) | 0.89 | 1.01 (0.87-1.17) | 0.84 |
| Moderate-Severe | 0.76 (0.16–3.64) | 0.66 | 0.26 (0.03–2.09) | 0.26 | 0.87 (0.70-1.09) | 0.24 |
| **Anxiety (ref=Minimal: 0-4)** |  |  |  |  |  |  |
| Mild | 1.36 (0.58-3.23) | 0.47 | 0.80 (0.34-1.87) | 0.61 | 0.98 (0.84-1.14) | 0.62 |
| Moderate to Severe | 0.36 (0.05- 2.84) | 0.33 | 0.33 (0.04-2.61) | 0.29 | 0.45 (0.10-2.01) | 0.29 |
| **TB Knowledge** |  |  |  |  |  |  |
| High | 0.80 (0.39-1.78) | 0.57 | 0.82 (0.40-1.55) | 0.48 | 0.82 (0.40-1.67) | 0.56 |
| **HIV Stigma [Median (IQR)]** | 0.98 (0.93-1.03) | 0.80 | 1.01 (0.94-1.08) | 0.83 | 1.02 (0.90-1.17) | 0.22 |
| **TB Stigma [Median (IQR)]** |  |  |  |  |  |  |
| Isolation | 1.02 (0.84-1.24) | 0.88 | 1.03 (0.86-1.24) | 0.72 | 1.02 (0.90-1.17) | 0.71 |
| Disclosure | 0.99 (0.84-1.16) | 0.87 | 1.01 (0.86-1.19) | 0.86 | 1.03 (0.93-1.13) | 0.57 |
| **Medical Mistrust (ref=Low)** |  |  |  |  |  |  |
| Medium | 1.25 (0.47-3.32) | 0.66 | 1.4 (0.62-3.18) | 0.42 | 1.34 (0.69-2.6) | 0.39 |
| High | 1.38 (0.48-3.92) | 0.55 | 0.66 (0.25-1.74) | 0.40 | 0.94 (0.45-1.97) | 0.87 |

<sup>a</sup> Odds ratios are from a multinomial logistic regression
<sup>b</sup> Odds ratios are from a logistic regression

**Table 4.** Logistic regression analysis of associations between female participants’ characteristics and suboptimal treatment engagement trajectory group, adjusted for age and education level.

| Characteristics | Suboptimal from Initiation |  |
| --- | --- | --- |
|  | aOR | p-value |
| **Relationship Status** |  |  |
| In a relationship | 0.42 (0.11–1.58) | 0.19 |
| **Lives Alone** |  |  |
| No | 0.95 (0.25–3.65) | 0.94 |
| **Live with Children** |  |  |
| Yes | 2.23 (0.27–18.38) | 0.45 |
| **Employment Status** |  |  |
| Unemployed | 1.22 (0.32–4.64) | 0.76 |
| **Monthly Household Income (ref=<R2000)** |  |  |
| R2000–R5000 | 1.25 (0.42–3.69) | 0.68 |
| >R5000 | 1.30 (0.13–13.30) | 0.83 |
| **Social Support (ref=High)** |  |  |
| Low | 1.53 (0.16–14.74) | 0.71 |
| Moderate | 1.09 (0.37–3.16) | 0.88 |
| **Ever had TB (ref=Never)** |  |  |
| Yes, less than 2 years ago | **5.22 (1.11–24.44)** | **0.04** |
| Yes, more than 2 years ago | 2.99 (0.96–9.32) | 0.06 |
| **HIV Status (ref=Positive)** |  |  |
| Negative | **0.33 (0.09–1.25)** | **0.10** |
| Unknown | 0.97 (0.10–9.71) | 0.98 |
| **Depression (ref=Minimal: 0-4)** |  |  |
| Mild | 1.52 (0.34–6.87) | 0.58 |
| Moderate | 1.47 (0.45–4.77) | 0.52 |
| Moderate-Severe | 1.88 (0.41–8.74) | 0.42 |
| **Anxiety (ref=Minimal: 0-4)** |  |  |
| Mild (5-9) | 2.15 (0.72–6.38) | 0.17 |
| Moderate to Severe ($\geq 10$) | 1.31 (0.32–5.42) | 0.71 |
| **TB Knowledge** |  |  |
| High | 1.53 (0.56–4.13) | 0.40 |
| **HIV Stigma [Median (IQR)]** | 1.02 (0.85–1.22) | 0.82 |
| **TB Stigma [Median (IQR)]** |  |  |
| Isolation | 0.97 (0.81–1.16) | 0.73 |
| Disclosure | 1.14 (0.97–1.33) | 0.12 |
| **Medical Mistrust (ref = Low)** |  |  |
| Medium | 1.59 (0.49–5.11) | 0.43 |
| High | 0.98 (0.24–4.06) | 0.98 |

## DISCUSSION

Latent-class growth modeling of treatment refill data among those completing treatment identified three distinct engagement patterns characterized by consistent engagement in care (70% of overall cohort), suboptimal engagement after two months (6% of overall cohort), and suboptimal engagement from treatment initiation (7% of overall cohort). The inclusion of these suboptimal engagement classes more than doubles the proportion of individuals in the cohort at risk for poor outcomes from 11% to 24%. Sex-stratified models showed that while both men and women shared two similar trajectories, men exhibited a distinct third trajectory not observed among women, underscoring important gender differences in engagement patterns. These findings indicate that standard binary metrics of treatment completion substantially underestimate the true burden of risk and highlight the need for gender-responsive interventions to support sustained engagement in care.[7–9,27]

Our findings highlight key limitations in how treatment success is programmatically defined. Evidence suggests that missing more than 10% of doses meaningfully increases the risk of poor outcomes.[8,9] In our study, patients in the “consistent engagement” class stayed below this threshold, while those in suboptimal classes routinely exceeded it. Furthermore, even though few individuals in our cohort met the programmatic definition of loss to follow-up (>60 consecutive days), many accumulated more than 60 total missed refill days. By focusing narrowly on completion status and disregarding cumulative engagement patterns, current monitoring frameworks and indicators may underestimate the proportion of individuals at risk for relapse, resistance, and other adverse outcomes. A consistent predictor of suboptimal engagement was having had TB within the past two years. This highlights persistent vulnerabilities, likely rooted in social, economic, behavioral, or health system challenges, and underscores the need for intensified counseling or support interventions in this group that may need to be adjustable to treatment stage.[13,28,29]

Gender-disaggregated analyses revealed two common trajectories among men and women (i.e., consistent engagement and suboptimal engagement from initiation), and that a history of TB in the past two years was associated with higher odds of suboptimal engagement among both men and women. However, men displayed a distinct “suboptimal engagement after two months” trajectory not observed among women. This trajectory may reflect men’s gendered social and economic pressures, such as the need to return to work or resume normative roles tied to masculinity, which could drive poor engagement in care once they begin to feel better, which is usually concomitant with the end of intensive phase of treatment.[30–33] Among men, HIV-negative status was a strong predictor of poor engagement from the start of treatment. This likely reflects gendered-barriers to men’s engagement in health services and lower baseline engagement with the health system, as men living without HIV may have fewer prior touch points with care and less familiarity navigating services compared to their peers living with HIV.[34,35]

These findings carry important implications for efforts to shorten TB treatment regimens. While four-month regimens are promoted as a solution to adherence challenges,[36,37] our results caution that shorter regiments may not resolve underlaying patters of suboptimal engagement. This is especially notable among the subgroup of individuals–predominantly men–who demonstrate suboptimal engagement in care once they begin to feel better. In this context, the concept of regimen “forgiveness,” or the capacity to tolerate clustered or intermittent missed doses without compromising cure, may be more consequential than duration itself.[38] Optimizing treatment strategies will require both biomedical innovation to enhance regimen forgiveness and patient-centered approaches to sustain engagement.

Applying latent-class growth modeling only among individuals defined as successfully completing treatment allowed us to identify heterogeneity in care engagement patterns that would have been otherwise masked/obscured by binary programmatic outcomes.[39] This approach offers a more nuanced understanding of when suboptimal engagement patterns occur, and risk factors associated with different patterns of engagement; putting forth a testable hypothesis for why individuals with a previous history of TB that “successfully” completed treatment may still be at risk of poor outcomes. In addition, stratifying models by gender provided new insights into sex-specific risks for suboptimal engagement in care that would otherwise be obscured in pooled analyses.[40] At the same time, limitations should be acknowledged. Confidence intervals around some associations were wide, reflecting modest sample size, and limiting our power to detect additional gender-specific predictors; larger cohorts may clarify whether non-significant associations with large effect sizes represent true underlying risks. Additionally, although our models illuminate engagement trajectories, they cannot establish causality, and the determinants of disengagement likely extend beyond the variables available in this dataset.

Engagement in TB care is more dynamic than current programmatic metrics capture. Trajectory modeling reveals heterogeneity that has direct implications for intervention timing and design. Gendered patterns, recent TB history, and heightened risk among HIV-negative men highlight opportunities for differentiated programming. Beyond research, trajectory modeling can serve as a practical analytic tool for TB programs by tracking shifts in the proportion of patients in suboptimal engagement classes and whether engagement curves are flattening toward consistent adherence. Such application can offer new insights into program quality and guiding strategies to improve outcomes. Future studies should validate these findings in larger cohorts and further evaluate the utility of trajectory-based monitoring as a complement to existing metrics.

## Data Availability

All data produced in the present study are available upon reasonable request to the authors

## NOTES

### Supplementary Data

Supplementary materials are available at Clinical Infectious Diseases online. Consisting of data provided by the authors to benefit the reader, the posted materials are not copyedited and are the sole responsibility of the authors, so questions or comments should be addressed to the corresponding author.

### Acknowledgements and Author Contributions

We thank the collaborating healthcare facilities, the Eastern Cape Provincial and the Buffalo City Metropolitan Department of Health for their support and permission. We give special thanks to Ralph Mawarire (Project Manager) and the implementation team/field workers for their input to this study. We acknowledge the participants for their time and contributions to this study.

AMM and JD conceptualized the study, developed the methodology, acquired study funding and provided study oversight. DB, KF and LDV managed field staff, study implementation, and project administration activities. EA conducted the formal analysis, including the visualization and data preparation. AMM and EA wrote the initial manuscript. JD, DB, KF, SC and LDV edited and reviewed all manuscript drafts. All authors have reviewed and approve the final submitted manuscript draft.

### Disclaimer

The content presented here is solely the responsibility of the authors and does not necessarily represent the official views of the National Institutes of Health (NIH).

### Data Sharing Statement

Data are available upon request.

## SUPPLEMENTAL TABLES

**Table S1:** Model Evaluation Metrics.

| Model | Number of Classes | AIC | BIC | Entropy |
| --- | --- | --- | --- | --- |
| **Overall Cohort** |  |  |  |  |
| Linear | 2 | 29513.14 | 29547.59 | 0.001 |
| Linear | 3 | 29292.99 | 29340.36 | 0.496 |
| Linear | 4 | 29298.99 | 29359.28 | 0.402 |
| Quadratic | 2 | 26593.70 | 26641.07 | 0.948 |
| Quadratic | 3 | **26298.14** | **26362.74** | 0.940 |
| Quadratic | 4 | 26306.14 | 26387.96 | 0.573 |
| Cubic | 2 | 26467.96 | 26515.33 | **0.955** |
| Cubic | 3 | 26475.96 | 26540.56 | 0.478 |
| Cubic | 4 | 26322.81 | 26404.63 | 0.608 |
| **Male Cohort** |  |  |  |  |
| Linear | 2 | 19976.90 | 20008.13 | 0.921 |
| Linear | 3 | 19982.90 | 20025.86 | 0.406 |
| Linear | 4 | 19988.90 | 20043.58 | 0.443 |
| Quadratic | 2 | 18403.11 | 18446.06 | 0.942 |
| Quadratic | 3 | **18218.89** | **18277.47** | 0.928 |
| Quadratic | 4 | 18226.89 | 18301.10 | 0.536 |
| Cubic | 2 | 18304.87 | 18347.83 | **0.943** |
| Cubic | 3 | 18221.66 | 18280.25 | 0.916 |
| Cubic | 4 | 18229.66 | 18303.87 | 0.552 |
| **Female Cohort** |  |  |  |  |
| Linear | 2 | 9115.42 | 9141.01 | **0.989** |
| Linear | 3 | 9121.42 | 9156.61 | 0.398 |
| Linear | 4 | 9127.42 | 9172.20 | 0.747 |
| Quadratic | 2 | **7720.05** | **7634.23** | 0.958 |
| Quadratic | 3 | 7761.79 | 7809.77 | 0.977 |
| Quadratic | 4 | 7763.99 | 7724.77 | 0.959 |
| Cubic | 2 | 7877.12 | 7912.30 | 0.979 |
| Cubic | 3 | 7791.15 | 7839.13 | 0.966 |
| Cubic | 4 | 7799.15 | 7859.92 | 0.540 |

**Table S2.**
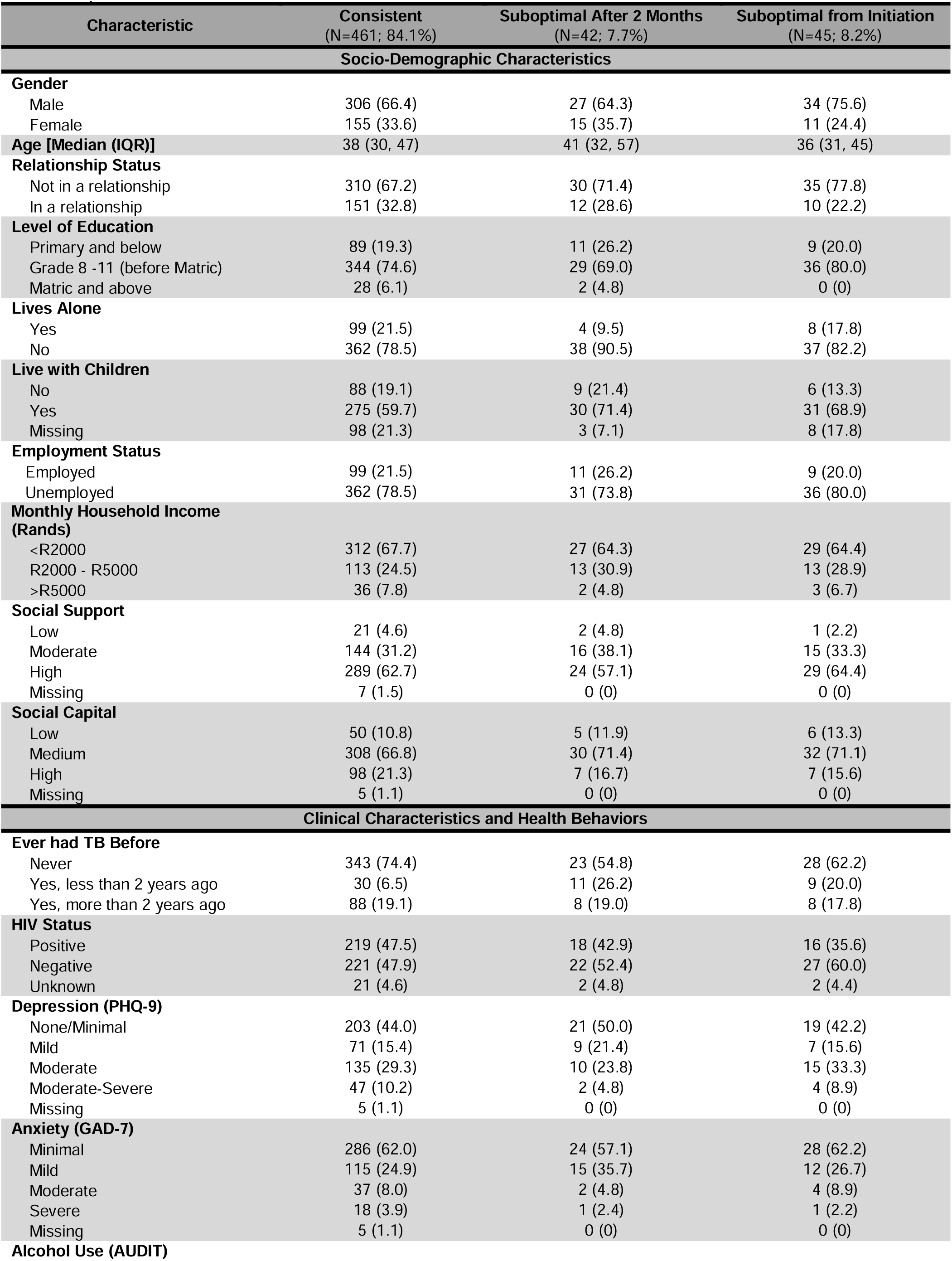
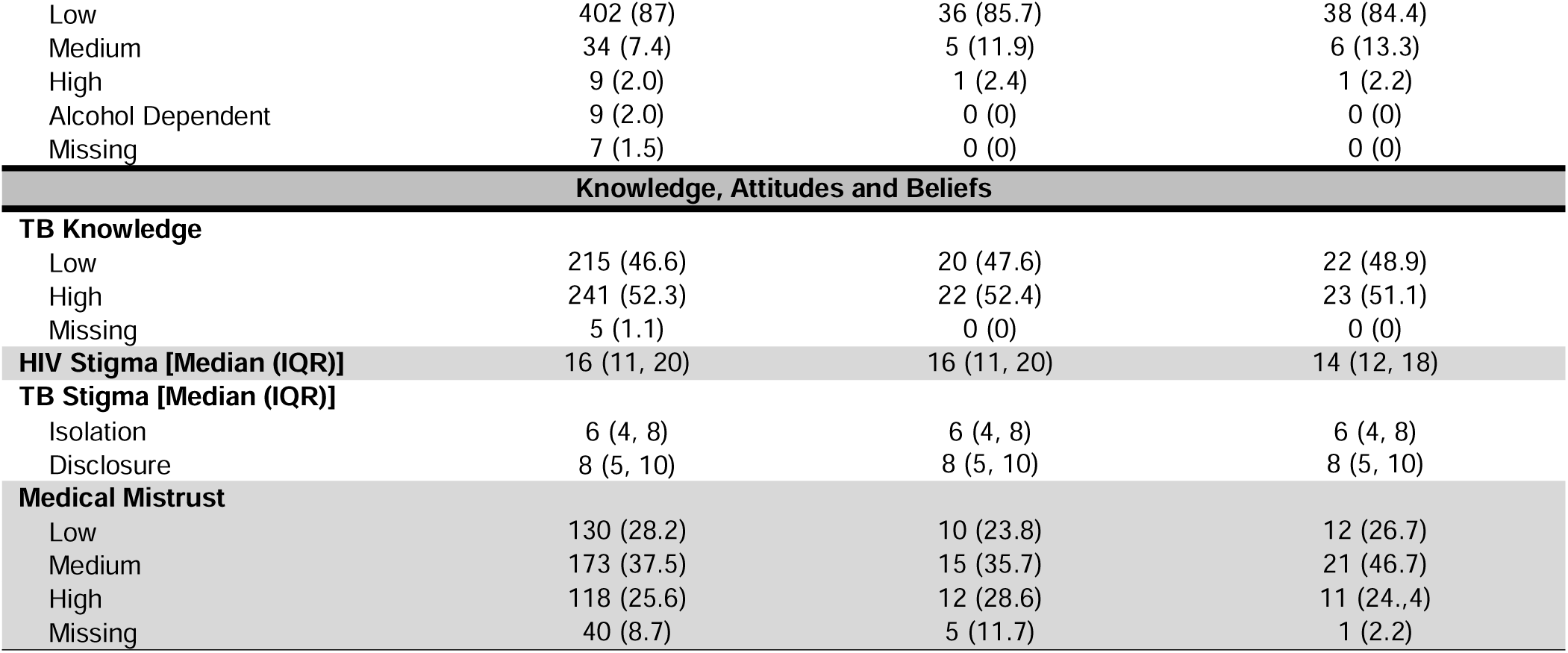
Descriptive Statistics of Study Participant’s Characteristics Stratified by Trajectory Group (All data are presented as N (%) unless stated otherwise)

**Table S3.** Characteristics of Male Study Participants Stratified by Trajectory Group (All data are presented as N (%) unless stated otherwise)

| Characteristics | Consistent<br>(N=305; 83.1%) | Suboptimal After 2 Months<br>(N=27; 7.4%) | Suboptimal From Onset<br>(N=35; 9.5%) |
| --- | --- | --- | --- |
| **Socio-Demographic Characteristics** |  |  |  |
| **Age** | 39 (31, 47) | 41 (32, 47) | 36 (31, 49) |
| **Relationship Status** |  |  |  |
| Not in a relationship | 203 (66.6) | 21 (77.8) | 26 (74.3) |
| In a relationship | 102 (33.4) | 6 (22.2) | 9 (25.7) |
| **Level of Education** |  |  |  |
| Primary and below | 71 (23.2) | 5 (18.5) | 6 (17.1) |
| Grade 8-11 (before Matric) | 220 (72.1) | 21 (77.8) | 29 (82.9) |
| Matric and above | 14 (4.6) | 1 (3.7) | 0 (0) |
| **Lives Alone** |  |  |  |
| Yes | 75 (24.6) | 4 (14.8) | 6 (17.1) |
| No | 230 (75.4) | 23 (85.2) | 29 (82.9) |
| **Live with Children** |  |  |  |
| No | 71 (23.3) | 6 (22.2) | 5 (14.3) |
| Yes | 160 (52.5) | 18 (66.7) | 24 (68.6) |
| Missing | 74 (24.2) | 3 (11.1) | 6 (17.1) |
| **Employment Status** |  |  |  |
| Employed | 69 (22.7) | 7 (25.9) | 8 (22.9) |
| Unemployed | 236 (77.3) | 20 (74.1) | 27 (77.1) |
| **Household Income in Rands** |  |  |  |
| <R2000 | 210 (68.9) | 19 (70.4) | 22 (63) |
| R2000 - R5000 | 70 (22.9) | 6 (22.2) | 11 (31) |
| >R5000 | 25 (8.2) | 2 (7.4) | 2 (5.7) |
| **Social Support** |  |  |  |
| Low | 14 (4.6) | 2 (7.4) | 0 (0) |
| Moderate | 99 (32.5) | 11 (40.7) | 12 (34.3) |
| High | 187 (61.3) | 14 (51.9) | 23 (65.7) |
| Missing | 5 (1.6) | 0 (0) | 0 (0) |
| **Social Capital** |  |  |  |
| Low | 32 (10.5) | 2 (7.4) | 4 (11.4) |
| Medium | 197 (64.6) | 20 (74.1) | 27 (77.1) |
| High | 71 (23.3) | 5 (18.5) | 4 (11.4) |
| Missing | 5 (1.6) | 0 (0) | 0 (0) |
| **Clinical Characteristics and Health Behaviors** |  |  |  |
| **Ever had TB Before** |  |  |  |
| Never | 222 (72.8) | 12 (44.4) | 24 (68.6) |
| Yes, less than 2 years ago | 24 (7.9) | 9 (33.3) | 7 (20.0) |
| Yes, more than 2 years ago | 59 (19.3) | 6 (22.2) | 4 (11.4) |
| **HIV Status** |  |  |  |
| Positive | 122 (40.0) | 10 (37.0) | 7 (20.0) |
| Negative | 169 (55.4) | 15 (55.6) | 26 (74.3) |
| Unknown | 14 (4.6) | 2 (7.4) | 2 (5.7) |
| **Depression (PHQ-9)** |  |  |  |
| None/Minimal | 141 (46.2) | 13 (48.1) | 16 (45.7) |
| Mild | 44 (14.4) | 6 (22.2) | 7 (20) |
| Moderate | 85 (27.9) | 6 (22.2) | 11 (31.4) |
| Moderate-Severe | 30 (9.8) | 2 (7.4) | 1 (2.9) |
| Missing | 5 (1.6) | 0 (0) | 0 (0) |
| **Anxiety (GAD-7)** |  |  |  |
| Minimal | 192 (63.0) | 17 (63.0) | 25 (71.4) |
| Mild | 77 (25.2) | 9 (33.3) | 8 (22.9) |
| Moderate | 22 (7.2) | 1 (3.7) | 1 (2.9) |
| Severe | 9 (3.0) | 0 (0) | 1 (2.9) |
| Missing | 5 (1.6) | 0 (0) | 0 (0) |
| **Alcohol Use (AUDIT)** |  |  |  |
| Low | 259 (84.9) | 23 (85.2) | 30 (85.7) |
| Medium | 26 (8.5) | 4 (14.8) | 4 (11.4) |
| High | 7 (2.3) | 0 (0) | 1 (2.9) |
| Alcohol Dependent | 8 (2.6) | 0 (0) | 0 (0) |
| Missing | 5 (1.6) | 0 (0) | 0 (0) |
| **Knowledge, Attitudes and Beliefs** |  |  |  |
| **TB Knowledge** |  |  |  |
| Low | 132 (43.3) | 13 (48.1) | 17 (48.6) |
| High | 168 (55.1) | 14 (51.9) | 18 (51.4) |
| Missing | 5 (1.6) | 0 (0) | 0 (0) |
| **HIV Stigma [Median (IQR)]** | 16 (11, 20) | 14 (7, 17) | 16 (12, 21) |
| **TB Stigma [Median (IQR)]** |  |  |  |
| Isolation | 6 (4, 8) | 6 (4, 8) | 6 (5, 8) |
| Disclosure | 8 (5, 10) | 8 (4, 10) | 8 (6, 10) |
| **Medical Mistrust** |  |  |  |
| Low | 85 (27.9) | 5 (18.5) | 11 (31.4) |
| Medium | 111 (36.4) | 8 (29.6) | 15 (42.9) |
| High | 82 (26.8) | 10 (37.0) | 8 (22.9) |
| Missing | 27 (8.9) | 4 (14.8) | 1 (2.9) |

**Table S4.** Characteristics of Female Study Participants Stratified by Trajectory Group (All data are presented as N (%) unless stated otherwise)

| Characteristic | Consistent<br>(N=162; 89.5%)* | Suboptimal from Onset<br>(N=19; 10.5%) |
| --- | --- | --- |
| **Socio-Demographic Characteristics** |  |  |
| **Age** | 38 (30, 48) | 30 (25, 42) |
| **Relationship status** |  |  |
| Not in a relationship | 129 (79.6) | 13 (68.4) |
| In a relationship | 63 (38.9) | 6 (31.6) |
| **Level of Education** |  |  |
| Primary and below | 22 (13.6) | 5 (26.3) |
| Grade 8 -11 (before Matric) | 126 (77.8) | 13 (68.4) |
| Matric and above | 14 (8.6) | 1 (5.3) |
| **Lives Alone** |  |  |
| Yes | 23 (14.2) | 3 (15.8) |
| No | 139 (85.8) | 16 (84.2) |
| **Live with Children** |  |  |
| No | 20 (12.3) | 1 (5.3) |
| Yes | 119 (73.5) | 15 (78.9) |
| Missing | 23 (14.2) | 3 (15.8) |
| **Employment Status** |  |  |
| Employed | 32 (19.8) | 3 (15.8) |
| Unemployed | 130 (80.2) | 16 (84.2) |
| **Monthly Household Income (Rands)** |  |  |
| <R2000 | 105 (64.8) | 12 (63.2) |
| R2000 - R5000 | 46 (28.4) | 6 (31.6) |
| >R5000 | 11 (6.8) | 1 (5.3) |
| **Social Support** |  |  |
| Low | 7 (4.3) | 1 (5.3) |
| Moderate | 47 (29.0) | 6 (31.6) |
| High | 106 (65.4) | 12 (63.2) |
| Missing | 2 (1.2) | 0 (0) |
| **Social Capital** |  |  |
| Low | 19 (11.7) | 4 (21.1) |
| Medium | 114 (69.5) | 12 (63.2) |
| High | 29 (17.9) | 3 (15.8) |
| **Clinical Characteristics and Health Behaviors** |  |  |
| **Ever Had TB Before** |  |  |
| Never | 126 (77.8) | 10 (52.6) |
| Yes, less than 2 years ago | 7 (4.3) | 3 (15.8) |
| Yes, more than 2 years ago | 29 (17.9) | 6 (31.6) |
| **HIV Status** |  |  |
| Positive | 99 (61.1) | 15 (78.9) |
| Negative | 57 (35.2) | 3 (15.8) |
| Unknown | 6 (3.7) | 1 (5.3) |
| **Depression (PHQ-9)** |  |  |
| None/Minimal | 67 (41.4) | 6 (31.8) |
| Mild | 27 (16.7) | 3 (15.8) |
| Moderate | 51 (31.5) | 7 (36.8) |
| Moderate-Severe | 17 (10.5) | 3 (15.8) |
| **Anxiety (GAD-7)** |  |  |
| Minimal | 96 (59.2) | 8 (42.1) |
| Mild | 40 (24.7) | 8 (42.1) |
| Moderate | 16 (9.9) | 3 (15.8) |
| Severe | 10 (6.2) | 0 (0) |
| **Alcohol Use (AUDIT)** |  |  |
| Low | 149 (92.0) | 15 (78.9) |
| Medium | 8 (4.9) | 3 (15.8) |
| High | 3 (1.9) | 0 (0) |
| Alcohol Dependent | 0 (0) | 1 (5.3) |
| **Knowledge, Attitudes and Beliefs** |  |  |
| **TB Knowledge** |  |  |
| Low | 86 (53.1) | 9 (47.4) |
| High | 76 (46.9) | 10 (52.6) |
| **HIV Stigma [Median (IQR)]** | **16 (12, 21)** | **20 (12, 23)** |
| **TB Stigma [Median (IQR)]** |  |  |
| Isolation | 6 (4, 8) | 7 (4, 8) |
| Disclosure | 9 (5, 10) | 10 (7, 11) |
| **Medical Mistrust** |  |  |
| Low | 46 (28.4) | 5 (26.3) |
| Medium | 65 (40.1) | 10 (52.6) |
| High | 37 (22.8) | 4 (21.5) |
| Missing | 14 (8.6) | 0 (0) |

